# Improvements in Albuminuria Screening Among Individuals with Hypertension Associated with Electronic Health Record Clinical Decision Support Design Changes

**DOI:** 10.64898/2026.02.09.26345709

**Authors:** Waleed Zafar, Spencer Tavares, Yirui Hu, Lauren Brubaker, Jamie Green, Sneha Mehta, Morgan E. Grams, Alexander R. Chang

## Abstract

**Background:** Albuminuria is associated with increased risk of cardiovascular disease (CVD), heart failure, and progression of chronic kidney disease (CKD). Early detection of albuminuria, done through spot urine albumin creatinine ratio (UACR) testing, enables more accurate risk stratification and timely use of preventative therapies. It remains unacceptably low in the hypertension population.

**Methods:** We evaluated two EHR-embedded clinical decision support (CDS) strategies at Geisinger Health System in order to increase UACR testing in individuals with hypertension: an OurPractice Advisory (OPA) from Jan 2022 to Aug 2022; and a Health Maintenance Topic (HMT) in the Care Gaps section of Storyboard from Aug 2022 that continues to date. We evaluated UACR rates from 2020 to 2023 in Geisinger primary care and compared to a control group of healthcare systems in the Optum Labs Data Warehouse [OLDW]. Patients were excluded if they had UACR testing in the preceding 3 years, had diabetes or CKD, or were receiving palliative/hospice care.

**Results:** We included 58,876 individuals in Geisinger (mean age 59.4 years, 49.6% female) and 1,427,754 in OLDW (61.0 years, 49% female). UACR testing in Geisinger (2.97% in 2020; 2.8% in 2021; 9.7% in 2022; 17.5% in 2023) showed significant increase compared to the control health systems (2.08%, 2.26%, 3.35% and 3.40% respectively). Results were consistent after adjusting for age, sex and race.

**Conclusion:** OPA increased UACR testing ∼3-fold whereas the HMT was associated with further improvements (∼6-fold vs. baseline) among those with hypertension, suggesting an important role for CDS design in closing care gaps.

## Introduction

Albuminuria, an early marker of kidney disease, is also a key risk factor for incident cardiovascular disease (CVD)^1^ likely through shared pathological processes such as vascular endothelial damage, inflammation, activation of the renin-angiotensin-aldosterone system, and neurohormonal activation^2^. The close interplay between metabolic risk factors (i.e. obesity, diabetes mellitus [DM]), CKD and CVD has been recently recognized by the American Heart Association (AHA) in a scientific statement on cardiovascular-kidney-metabolic (CKM) health^3^. Screening for CKD (estimated glomerular filtration rate [eGFR] and albuminuria) is thus a critical step in assessing CKM health. Current guidelines recommend at least annual UACR testing for individuals with DM^4,5^ and more frequent for individuals with CKD^6^. While the European Society of Cardiology and European Society of Hypertension has recommended UACR testing at least once for those with hypertension since 2018^7^, the American Heart Association (AHA) just recently updated their Hypertension Guidelines in 2025 Hypertension to recommend UACR testing for individuals with hypertension “at the time of diagnosis and possibly annually thereafter”^8^. Timely diagnosis of albuminuria can also help risk stratify patients in need of more intensive therapy, which is underscored by the inclusion of albuminuria in the AHA’s Predicting Risk of CVD EVENTs (PREVENT) calculator^9^.

Unfortunately, rates of albuminuria screening remain unacceptably low among high-risk patients, especially among those with hypertension^10–12^. In an analysis of 2.3 million adults, mostly between 2011 to 2020, mean albuminuria testing rates were only 4.1% (range 1.3% to 20.7%) for individuals with hypertension without DM, and 35.1% (range 12.3% to 74.5%) for individuals with DM^12^. The landscape for effective treatment of albuminuria has changed dramatically with the advent of cardiorenal protective medications, including sodium-glucose cotransporter-2 inhibitors (SGLT2i) and for those also with DM, glucagon-like peptide-1 (GLP-1) receptor agonists and nonsteroidal mineralocorticoid receptor antagonists (nsMRAs)^6^. Addressing albuminuria testing is the first step to address other significant care-gaps in the management of CKD as detection of new albuminuria is associated with 3-fold higher prescription of renin angiotensin aldosterone system (RAAS) inhibitors^12^.

Barriers to albuminuria screening in primary care remain underexplored but include providers’ perceived lack of impact on management, competing demands on time, lack of familiarity with guidelines^13^ and difficulty obtaining urine samples in the primary care office^14^. System-level changes may help overcome these barriers to improve albuminuria testing in primary care ^15^. For example, clinical decision support (CDS) has been identified as an important facilitator for improving CKD management by primary care providers.^16 17^ Well-designed CDS built into electronic health records (EHR) can link patient-specific data with evidence-based knowledge and case-specific messages to optimize clinical care.

In this study, we assessed albuminuria screening rates associated with two different CDS designs at a large healthcare system and controlled for temporal changes, by comparing results to data in the Optum Labs Data Warehouse (OLDW), a very large, longitudinal, real-world dataset with deidentified administrative claims and EHR data ^18^.

## Methods

### Study Population

This study included adult individuals receiving primary care at Geisinger, a large, mostly rural health system in central and northeast Pennsylvania providing care to approximately 1.2 million people. As an early adopter of EHR using Epic^®^ since 1996, Geisinger has leveraged high-quality clinical data assets to facilitate innovation, research, and quality improvement. Geisinger has a strong clinical informatics team including a clinical informatics fellowship training program, with health professionals across primary care, specialties, and population health, enabling the rapid testing and deployment of effective clinical decision support tools.^19,20^ For a comparator group, we created a control group using data from the OLDW,^18^ a deidentified, longitudinal health information database of administrative claims and EHR data representing a wide cross-section of ages and geographical regions representing many large US health systems.

Eligible population were all adults seen in primary care clinics who had a diagnosis of hypertension within the 2 years prior to the clinic visit and had not had a UACR test done in the previous three years. This criterion was used to represent patients with established diagnosis of hypertension who should receive UACR screening. Because there were already existing CDS to test UACR annually in patients with DM or CKD, we excluded individuals if they had a concurrent diagnosis of DM, CKD or end stage kidney disease (ESKD), HbA1c≥6.5%, or eGFR<60 twice during the study period. Additionally, we excluded individuals enrolled in palliative care or hospice care. This study was approved by the Geisinger Institutional Review Board.

### CDS Design of Albuminuria for Hypertension Our Practice Advisory (OPA)

After discussion amongst stakeholders including primary care providers, quality leaders, and clinical informatics, we developed an OPA within the Epic EHR at Geisinger starting in Jan 2022 alerting providers to order UACR for individuals with hypertension who had not completed testing in the preceding 3 years (Figure 1A). The alert was designed to be non-interruptive and utilized a passive trigger to display in the Best Practice navigator section of the chart. The OPA was restricted to primary care office visits with physician or advanced practice providers. Advisory criteria included adult patients with hypertension, no available UACR result in the past 3 years, and no active UACR order. The OPA displayed the action to place a UACR order. Providers could choose to place the order, acknowledge reason to defer the OPA (e.g. “not clinically appropriate”), or could ignore the OPA altogether. This built on similar OPA for DM and CKD but targeting specifically individuals with hypertension.

**Figure 1.**
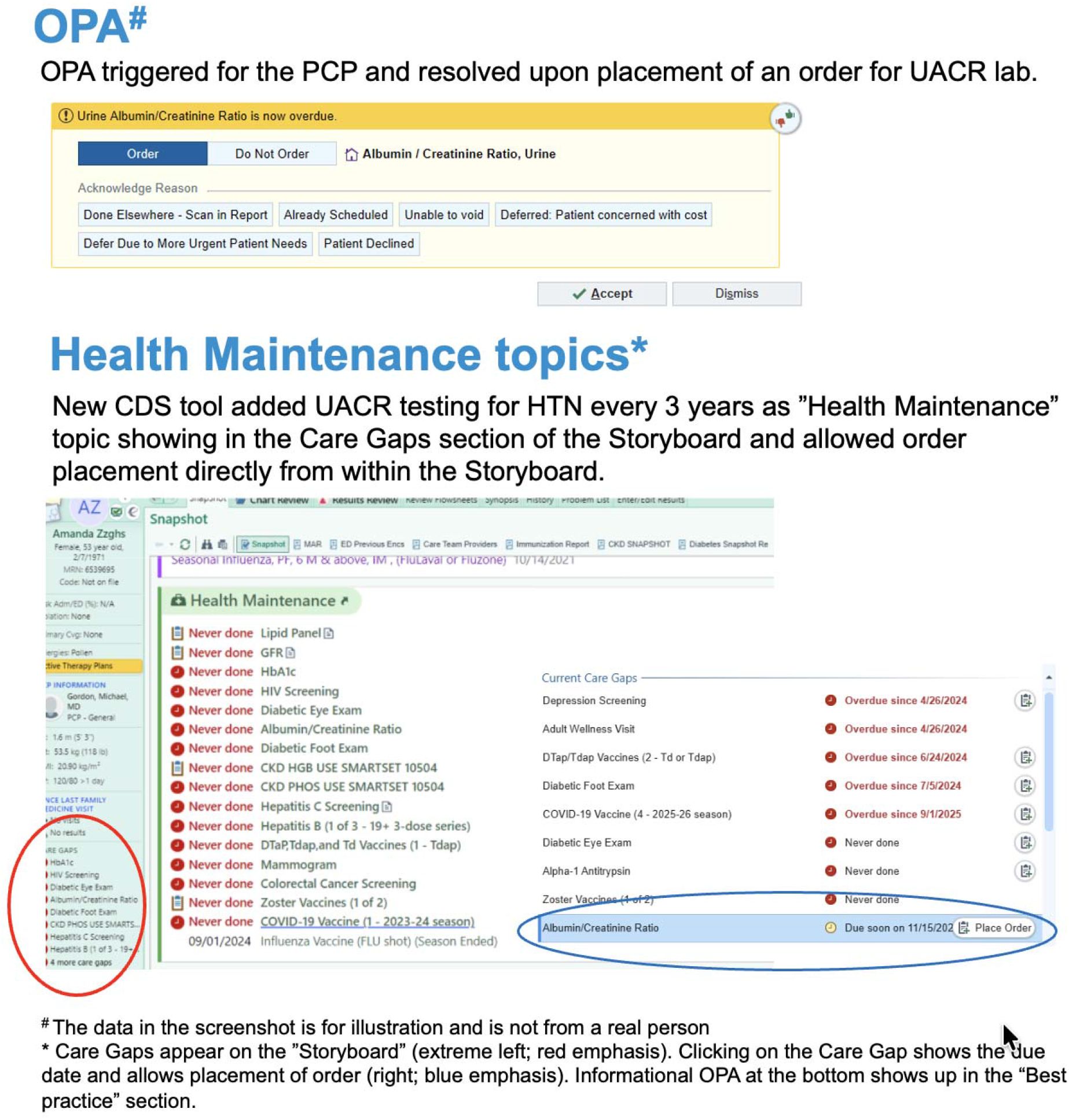
Example of OurPractice Alert (OPA) and Health Maintenance topic

### Storyboard Health Maintenance topic

In light of a system-wide feedback and literature suggesting that alerts were seen as a nuisance and could be associated with clinician alert fatigue and reduced adherence, the OPA was replaced as a more user-friendly “Health Maintenance Topic” showing up in the Care Gaps section of “Storyboard” within the Epic^®^ EHR (Verona, WI: Epic Systems; 2025). Clicking on the Care Gap showed the due date and allowed placement of the order directly from within the Storyboard. The UACR health maintenance topic used the same inclusion criteria as the OPA and utilized single-click UACR ordering, but health maintenance topics do not include acknowledgement reasons.

### Outcomes

The time period of the OPA was from 1/4/2022 to 8/16/2022. To measure the effect of switch to the “Storyboard” health maintenance topic, we used identical time periods (1/4/2023 to 8/16/2023) to control for any possible seasonality. To obtain baseline prior to implementation of OPA, we defined two time periods (1/4/2020 to 8/16/2020 and 1/4/2021 to 8/16/2021) prior to the start of OPA. The primary outcome was UACR test completion during each of these time periods.

### Statistical Analyses

Baseline characteristics were summarized for each time-period. Differences in screening rates by various sociodemographic factors were assessed (ANOVA for continuous variables and Chi-square tests for categorical variables). Multinomial logistic regressions were performed to evaluate the association between the OPA time-period (1/4/2022 to 8/16/2022) and the storyboard health maintenance topic time-period (1/4/2023 to 8/16/2023), compared to the baseline time-period (1/4/2020 to 8/16/2020) to estimate odds ratio (OR) with corresponding 95% confidence interval (CI). Adjustments were made for age, sex, race, and ethnicity. Each individual’s data was included only once with the earliest possible qualifying time-period meeting inclusion/exclusion criteria. Thus, individuals were not included in subsequent time periods to make time periods more comparable. Separate analyses were conducted for Geisinger and for the OLDW datasets using the same methods. For our analyses, the total number of included patients in Geisinger were 58,876 and in OLDW were 1,427,754. All analyses were performed in Rstudio^21^.

## Results

Baseline characteristics across each of the four time periods in Geisinger system and OLDW dataset are shown in Table 1. Geisinger population contained significantly more individuals self-identifying as white (∼94%) compared to OLDW data (64% to 72%). The average age in OLDW cohort across four time periods was higher in later years (2020 59 years; 2023 61 years) while it was lower in later years in the Geisinger cohort (2020 60 years; 2023 58 years), possibly because of increased testing in prior time periods leaving younger individuals untested in the subsequent cohort.

**Table 1:** Demographic characteristics of participants within Geisinger system and in the Optum Labs Data Warehouse (OLDW) dataset.

|  | Geisinger |  |  |  | OLDW |  |  |  |
| --- | --- | --- | --- | --- | --- | --- | --- | --- |
|  | 2020 | 2021 | 2022 | 2023 | 2020 | 2021 | 2022 | 2023 |
| **Age; Mean (SD)** | 60.02<br>(13.42) | 60.19<br>(13.57) | 59.92<br>(13.75) | 57.6<br>(14.22) | 59<br>(15) | 60<br>(15) | 61<br>(15) | 61<br>(15) |
| **Female; N (%)** | 6,937<br>(50.0%) | 8,096<br>(49.5%) | 8,578<br>(49.4%) | 5,614<br>(49.6%) | 248,852<br>(48%) | 236,269<br>(49%) | 122,974<br>(49%) | 84,893<br>(48%) |
| **Race /**<br>**Ethnicity; N (%)** |  |  |  |  |  |  |  |  |
| **White** | 13,105<br>(94.5%) | 15,433<br>(94.4%) | 16,393<br>(94.5%) | 10,623<br>(93.9%) | 370,624<br>(72%) | 340,028<br>(71%) | 168,667<br>(67%) | 113,186<br>(64%) |
| **Black** | 445<br>(3.2%) | 527<br>(3.2%) | 530<br>(3.1%) | 355<br>(3.1%) | 68,362<br>(13%) | 62,168<br>(13%) | 35,044<br>(14%) | 25,092<br>(14%) |
| **Hispanic\*** | 364<br>(2.6%) | 506<br>(3.1%) | 621<br>(3.6%) | 484<br>(4.3%) | 26,859<br>(5.2%) | 25,518<br>(5.3%) | 15,919<br>(6.3%) | 10,986<br>(6.2%) |
| **Asian** | 121<br>(0.9%) | 153<br>(0.9%) | 150<br>(0.9%) | 79<br>(0.7%) | 11,737<br>(2.3%) | 13,225<br>(2.7%) | 7,734<br>(3.1%) | 5,253<br>(3.0%) |
| **Other** | 190<br>(1.4%) | 238<br>(1.5%) | 278<br>(1.6%) | 256<br>(2.3%) | 38,646<br>(7.5%) | 40,724<br>(8.5%) | 25,935<br>(10%) | 22,047<br>(12%) |
| * Race and Hispanic ethnicity were treated separately in Geisinger data; therefore, column percentages may not sum to 100%. |  |  |  |  |  |  |  |  |

Figure 2 shows the main outcome across four time periods. Within Geisinger, proportion of individuals with completed UACR test was 3.0% and 2.8% in the two time periods before the institution of OPA and rose significantly higher to 9.7% during the OPA period. It further increased to 17.5% when the switch was made to the storyboard/HMT. After adjustment for age, sex, race, and ethnicity, OR for albuminuria testing at Geisinger were 3.54 (95% CI: 3.17, 3.96; p<0.001) in 2022 and 7.11 (95% CI: 6.38, 7.94; p<0.001) in 2023, compared to 2020. In comparison, the proportion completing UACR testing in OLDW increased minimally from 2020 (2.08%) to 2021 (2.26%), 2022 (3.35%), and 2023 (3.40%).

**Figure 2.**
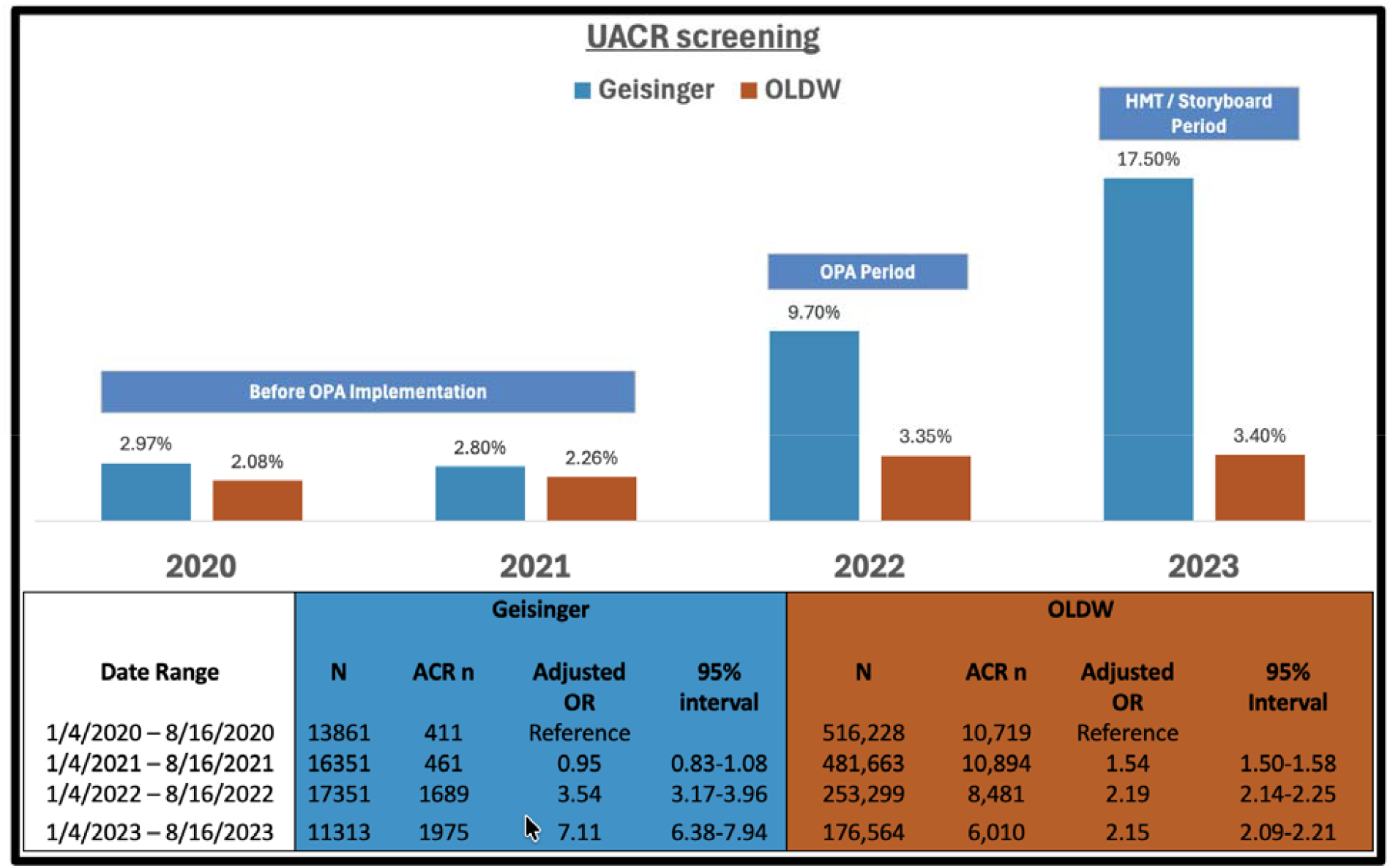
Change in UACR screening rates over time and between Geisinger and Optum Lab Data Warehouse (OLDW) data

## Discussion

In this study we evaluate the impact of CDS changes prompting PCPs to order UACR testing for individuals with hypertension (without DM or CKD) in a large regional health system. By comparing to a large national cohort spanning multiple health systems, we demonstrate that the type of CDS change can have a large impact on screening for albuminuria. After the implementation of an OPA at Geisinger, UACR testing increased ∼3-fold while after switching to a more user-friendly CDS, there was further improvement in UACR testing (∼6-fold vs. baseline) among patients with hypertension without diabetes or CKD. In comparison, there was minimal increase in UACR testing within OLDW health systems in the hypertension population.

Our study is one of the first to highlight that displaying a health screening need as a Health Maintenance topic within Epic’s Storyboard can substantially improve screening rates compared to a passive OPA embedded in the visit navigator. While both tools were non-interruptive, the Health Maintenance topic offered a more prominent and persistent visual cue by integrating care gaps directly into the Storyboard as a fixed, left-hand column visible throughout the clinical workflow. This design likely enhanced provider awareness and actionability by presenting albuminuria screening alongside other preventative care needs in a centralized, patient-centric format. In contrast, the OPA, although non-interruptive, was less visible and required deliberate navigation to access, which may have limited its effectiveness. The improved performance of the Storyboard tool underscores the importance of strategic CDS placement within the EHR interface to support proactive care delivery without contributing to alert fatigue. Table 2 compares the advantages of Epic’s Health Maintenance module in Storyboard versus passive OPAs, highlighting differences in visibility, workflow integration, and support for preventative care.

**Table 2:** A Comparison of Storyboard Health Maintenance topic and OurPractice Advisory.

|  | Health Maintenance Topics in<br>Storyboard | OurPractice Advisory (OPA) |
| --- | --- | --- |
| Visibility | Persistently visible on the left sidebar during chart review | Requires navigation within the visit navigator; not always visible; may appear as dismissible temporary pop-up or one time alert |
| Interruptiveness | Passive, non-interruptive display | Non-interruptive in this study; Can be overlooked due to lower visibility |
| Workflow Integration | Integrated into routine workflow | Less integrated; may be missed if not actively searched for |
| Contextual Relevance | Presents a comprehensive view of care gaps across multiple conditions | Focused on a single condition or recommendation at a time; less adaptable |
| Actionability | Hover for details and click to initiate actions (e.g., place orders) directly | Requires manual navigation and initiation of actions |
| Alert Fatigue Risk | Low – passive display minimizes cognitive burden and alert fatigue | Moderate – Limited visibility and possible alert redundancy increase fatigue risk |
| Preventive Care | Strong – designed to support chronic condition management and preventive care | Limited – better suited for acute issues |
| Provider Engagement | Moderate – care gaps are clearly visible and actionable during chart review | Low – risk of being overlooked due to less prominent placement |
| Patient Engagement | Moderate - Potential to link to patient engagement tools (e.g. My Chart) | Limited - Generally provider-facing; limited patient engagement potential |

Our work also adds to the nascent but growing literature on how provider feedback can be incorporated into a careful re-design of CDS. For instance, a recent study sought to understand referral of CKD patients to Nephrology through interviews with PCPs that explored referral decision making process and the specific changes in patient’s status that would prompt referral to Nephrology^22^. This process was then used to design a CDS system to encourage timely referral of CKD patients to Nephrology. Among the needs identified by this study were those for suggestive prompts, accurate identification of patients, ability to see trends in BP and eGFR values, ability to access needed information from within the workflow, and receipt of CDS at the time of decision making^22^. Another study used interviews with physicians to develop a CDS encouraging use of RAAS blockers in hypertensive individuals^23^. That study showed how user feedback can be incorporated into the design of CDS tools in an iterative process through the use of group sessions to understand the context in which CDS tools are employed and individual think aloud sessions to fine tune their implementation^23^. Further studies are needed to determine whether these types of user-centered designs can positively impact clinical care.

While improving albuminuria screening rates is a key first step in identifying kidney and cardiovascular risk, few large-scale studies have been able to evaluate improvement in CKD-related care gaps in primary care as a result of changes in CDS design, and to date many such studies have been characterized by significant heterogeneity^24–35^. In an early successful CDS example, Litvin et al. developed a risk assessment tool prompting annual eGFR and UACR testing and a CKD patient registry^27^. This study found that albuminuria screening for individuals with diabetes or hypertension improved from baseline 21.5% to 59% at 24 months. However, the CDS had no impact on use of ACEi/ARB, screening for anemia, dyslipidemia, or avoidance of non-steroidal anti-inflammatory. In another well-designed quality improvement initiative by Park et al., patients enrolled in a CKD registry who did not have a UACR within past year had an automatic UACR ordered, and PCPs received an EHR alert regarding initiation of ACEi / ARB and CKD care education sessions were conducted in primary care clinics to reinforce UACR testing and management^29^. At 2-year follow up, UACR testing increased from 26.9% at baseline to 83% at 1^st^ year and 77% at 2^nd^ year (83.9% and 75.2% respectively among patients who did not have diabetes). However, there was no improvement in the use of ACEi / ARB, glycemia control, or blood pressure control^29^. These findings, while encouraging, highlight the need for further prospective studies to evaluate the effect of well-designed CDS to improve utilization of goal-directed medical therapies to improve cardiovascular and kidney outcomes.

Among the strengths of this study is the use of two different CDS designs that allowed for comparison of these two designs separately from the baseline as well as with each other. To control for temporal variations in UACR testing, we compared the effect of these design changes on albuminuria screening rates with a representative sample of US health systems in a large dataset containing longitudinal health information representing a diverse mixture of ages, ethnicities and geographical regions. These comparisons strengthen our conclusion that the changes in UACR screening were, in large part, prompted by the CDS design changes. There are a few limitations to this study. While this was not a randomized trial, the inclusion of temporal control as well as comparison with a large dataset provides confidence that the CDS changes had a major impact. Additional prospective studies would be helpful to see if our findings are scalable to other health systems. Lastly, we did not know specifics about similar CDS deployed at other U.S. health systems in OLDW data. Future work will need to examine health system-level factors associated with higher albuminuria screening uptake, as well as additional CDS interventions to improve uptake of goal-directed medical therapies once albuminuria is detected.

### Perspectives

This study demonstrates that the design of clinical decision support (CDS) tools can significantly improve uptake of albuminuria screening among individuals with hypertension. Even though urine albumin/creatinine ratio (UACR) testing is central to identifying early cardiovascular and kidney risk, it is infrequently done in routine practice in patients with hypertension. By evaluating two CDS strategies in a large health system, we show that more visible and persistent cues embedded in the clinical workflow yield much higher testing rates than best practice alert reminders. However, since only limited care gaps may be displayed in such a format, careful prioritization and collaboration is needed among population health scientists, health informatics specialists, primary care providers, and specialists to identify gaps that have the potential for greatest population health impact. Thoughtful leveraging of CDS tools may be a scalable strategy to close persistent gaps in the early identification, risk stratification, and management of cardiovascular-kidney-metabolic risk.

## Conclusions

Implementation of an OPA increased UACR testing ∼3-fold while a more user-friendly CDS was associated with further improvements in UACR testing (∼6-fold vs. baseline) among patients with hypertension without diabetes or CKD. The improved performance of the HMT CDS tool underscores the importance of strategic CDS placement within the EHR interface to support proactive care delivery without contributing to alert fatigue.

### Novelty and Relevance

#### What is New?

After the implementation of an EHR alert at a predominantly rural large health system, UACR testing increased among patients with hypertension (without diabetes or CKD). After incorporating feedback leading to a more user-friendly clinical decision support, there was further improvement in UACR testing. In comparison, there was minimal increase in UACR testing in comparator health systems during the same time frame.

#### What is Relevant?

Strategic employment of EHR-based clinical decision support (CDS) tools that are responsive to end-user feedback can help improve guideline-concordant screening and management gaps among individuals with hypertension.

#### Clinical/Pathophysiological Implications?

By leveraging well-designed CDS tools, earlier albuminuria detection can lead to earlier diagnosis of CKD and more aggressive intervention to reduce cardiovascular-kidney-metabolic risk.

## Data Availability

Data may be available upon reasonable request, pending appropriate data use agreement.

## Funding

The study was supported by NIDDK R01 DK115534.

## Notes

### Competing Interest Statement

The authors have declared no competing interest.

### Author Declarations

This study was approved by the Geisinger Institutional Review Board.

